# Epidemiologic Characteristics of COVID-19 in Guizhou, China

**DOI:** 10.1101/2020.03.01.20028944

**Authors:** Kaike Ping, Mingyu Lei, Yun Gou, Ying Tao, Yan Huang

**Affiliations:** Guizhou Center for Disease Control and Prevention, Guizhou, China

**Author notes:** Corresponding author. **Address for correspondence:** Y. Huang. Institute of Communicable Disease Prevention and Control, Guizhou Center for Disease Control and Prevention, Guizhou, China. Contributed equally to the article (Kaike Ping, Mingyu Lei).

## Abstract

At the end of 2019, a coronavirus disease 2019 (COVID-19) outbroke in Wuhan, China, and spread to Guizhou province on January of 2020. To acquire the epidemiologic characteristics of COVID-19 in Guizhou, China, we collected data on 162 laboratory-confirmed cases related to COVID-19. We described the demographic characteristics of the cases and estimated the incubation period, serial interval and basic reproduction number. With an estimation of 8 days incubation period and 6 days serial interval, our results indicate that there may exist infectiousness during the incubation period for 2019-nCoV. This increases the difficulty of screening or identifying cases related to COVID-19.

## Background

At the end of 2019, a Novel Coronavirus Infected Pneumonia (NCIP) outbroke in Wuhan, China. This coronavirus was named as 2019-Novel Coronavirus (2019-nCoV), and then renamed this disease caused by the virus strain from 2019-nCoV acute respiratory disease to “coronavirus disease 2019” (COVID-19) by World Health Organization (WHO) officially^0^. This incident caused a public health issue within China immediately, and spread to several countries over the world^2–5^.

Guizhou province is in Southwest of China, close (∼1,000 km) to Wuhan. With the increasing number of COVID-19 patients in mainland China, Guizhou detected its first confirmed case on January 21, and was officially confirmed by Chinese Center for Disease Control and Prevention (China CDC) on January 23. Before the confirmation of the first case, Guizhou CDC identified and screened suspected COVID-19 cases by “pneumonia of unknown etiology” surveillance mechanism^6^. After the confirmation, Guizhou CDC started to identify and screen every case with pneumonia.

Until February 16, over 140 laboratory-confirmed cases were officially reported by Guizhou Health Committee^7^. In this study, we analyzed the data on the laboratory-confirmed cases in Guizhou to demonstrate the epidemiologic characteristics of COVID-19.

## Methods

### Case Definitions

According to standard clinical guidelines, suspected case of COVID-19 was defined as the combination of clinical characteristics and epidemiologic histories. Clinical characteristics of suspected case must fit at least 2 of 3 following criteria:

“fever and/or symptoms in respiratory system; radiographic evidence of pneumonia; low or normal white-cell count or low lymphocyte count.” ^8^ Epidemiologic histories must fit at least 1 of 4 following criteria: “A history of traveling Hubei Province or other districts that has confirmed cases reported within 14 days of symptom onset; A history of contacting with patient who has fever or symptoms in respiratory system from Hubei Province or other districts that has confirmed cases reported within 14 days of symptom onset; Any person who has had a close contact with confirmed cases; cluster cases.”^8^ With a stricter definition, Guizhou Health Committee counted all cases that fit clinical characteristics criteria but not any criteria of epidemiologic histories described above as suspected cases. A confirmed case was defined as a case with respiratory specimens that tested positive for the 2019-nCoV by at least one of the following two methods: positive result by real-time reverse-transcription– polymerase-chain-reaction (RT-PCR) assay for 2019-nCoV or a genetic sequence that matches 2019-nCoV. Asymptomatic carrier (i.e. case that displays no signs or symptoms to 2019-nCoV) was not announced by Health Committee officially, but was included in this study.

### Sources of Data

Suspected case was screened by local hospital or local Center for Disease Control and Prevention (CDC). Once the patient was identified as confirmed case by laboratory, a joint field epidemiology team comprising members from Guizhou CDC and local CDC would open a detailed field investigation on demography information, epidemiologic histories, timelines of key events, and close contacts.

All data from epidemiological reports were inputted into standardized forms from technical protocols designed by China CDC. At least two team members independently review the full report of each case report to ensure that data is correctly input.

### Epidemiological investigation

Once a case was evaluated as suspected case, local CDC would finish an initial investigation report in 24 hours and collect respiratory specimens for centralized laboratory testing. When case was confirmed as positive to 2019-nCoV, province CDC would send a special epidemiology team to investigate with local CDC, in order to acquire more detailed information. Information was collected from infected individuals, family members, medical workers, close contacts, GPS-info, and CCTV camera, etc. Information included basic demography data, detailed life trace and all close contacts, clinical characteristics, exposure history, etc.

### Laboratory confirmation

With the guidelines of protocol by World Health Organization (WHO) ^9^, specimens of suspected cases were taken from whose upper and lower respiratory tract by professional medical worker from CDC or hospital. RNA was extracted and tested by RT-PCR with primers and probes for 2019-nCoV. Cross-reactivity with other known respiratory viruses and bacteria such as Influenza A (H1N1, H3N2, H5N1, or H7N9), Influenza B (Victoria or Yamagata), MERS-CoV, Adenovirus, etc. are also be tested.

### Statistical analysis

The onset date was defined as the self-reported first date to have symptoms related to 2019-nCoV or the date of visiting to clinical facilities if it was unavailable to acquire the accurate onset date, and as the confirmation date for confirmed cases without symptoms. As the date of infecting exposure for a given individual falls within a finite interval, an exposure window was clarified to denote this interval. Criteria of infecting exposure was described above in the **Case Definitions** section. Continuous variables were expressed as the means and standard deviations or medians, the interquartile range (IQR) equals the difference between 75th and 25th percentiles. Categorical variables were summarized as the counts and percentages in each category. The incubation period is defined as the delay from viral infection to the onset of illness^10^, which was estimated by fitting a parametric accelerated failure time model with log-normal distribution of cases with detailed exposure window data, and standard errors of parameters was computed using a 1,000 times bootstrap routine, this was performed by R package *coarseDataTools*^11^. The serial interval distribution (i.e. the duration between symptom onset of a primary/index case and symptom onset of its secondary cases) was fitted by a Weibull distribution with data from cluster events. The basic reproduction number (R0) was defined as the expected number of cases directly generated by one case in a population where all individuals are susceptible to infection, it was estimated by maximum likelihood estimation via R package *R0*^12^.

All analyses and statistical graphs were conducted with R software version 3.6.2 (R Foundation for Statistical Computing).

### Ethics approval

Data collection and analysis of cases and their close contacts were determined by Guizhou Health Commission of the People’s Republic of China. It is part of a continuing public health outbreak investigation and exempted from institutional review board assessment.

## Results

There were 759 cases in total. Among all patients, 144 (18.97%) cases were confirmed cases with symptoms to 2019-nCOV, 18 (2.37% of all 759 cases, 11.11% of all 162 confirmed cases) cases were confirmed as cases without symptoms, 597 (78.65%) cases were diagnosed as other diseases after clinical diagnosing or laboratory testing, 34 (4.47%) cases were suspected cases that were pending for laboratory testing. **Figure 1** shows the geographic distribution of confirmed cases throughout China and Guizhou. The median age of all confirmed cases was 37 years (ranges from 1 month to 89 years), 82 (50.62%) confirmed cases were male, 84 (51.85%) confirmed cases were exposed to Wuhan or other districts outside Guizhou, the detailed demographic data are shown in **Table 1**.

**Table 1.** Demographic Characteristics of Cases with COVID-19 in Guizhou

| Characteristics | Value |
| --- | --- |
| Sample size | 162 |
| Cases with symptoms | 144 |
| Cases without symptoms | 18 |
| Median age (range) — yr | 37 (1 month - 89) |
| Age group — no. (%) |  |
| <15 yr, | 16 (9.88%) |
| 15–44 yr, | 88 (54.33%) |
| 45–64 yr, | 46 (28.39%) |
| ≥65 yr | 12 (7.40%) |
| sex |  |
| Male | 82 (50.62%) |
| Female | 80 (49.38%) |
| Exposure info — no. (%) |  |
| Detailed exposure window | 93 (58.02%) |
| Only right bound of exposure window known | 54 (33.33%) |
| exposure window unknown | 4 (2.47%) |
| Onset date unknow | 11 (6.18%) |
| Exposure in Wuhan or other districts outside Guizhou— no. (%) | 84 (51.85%) |

**Figure 1.**
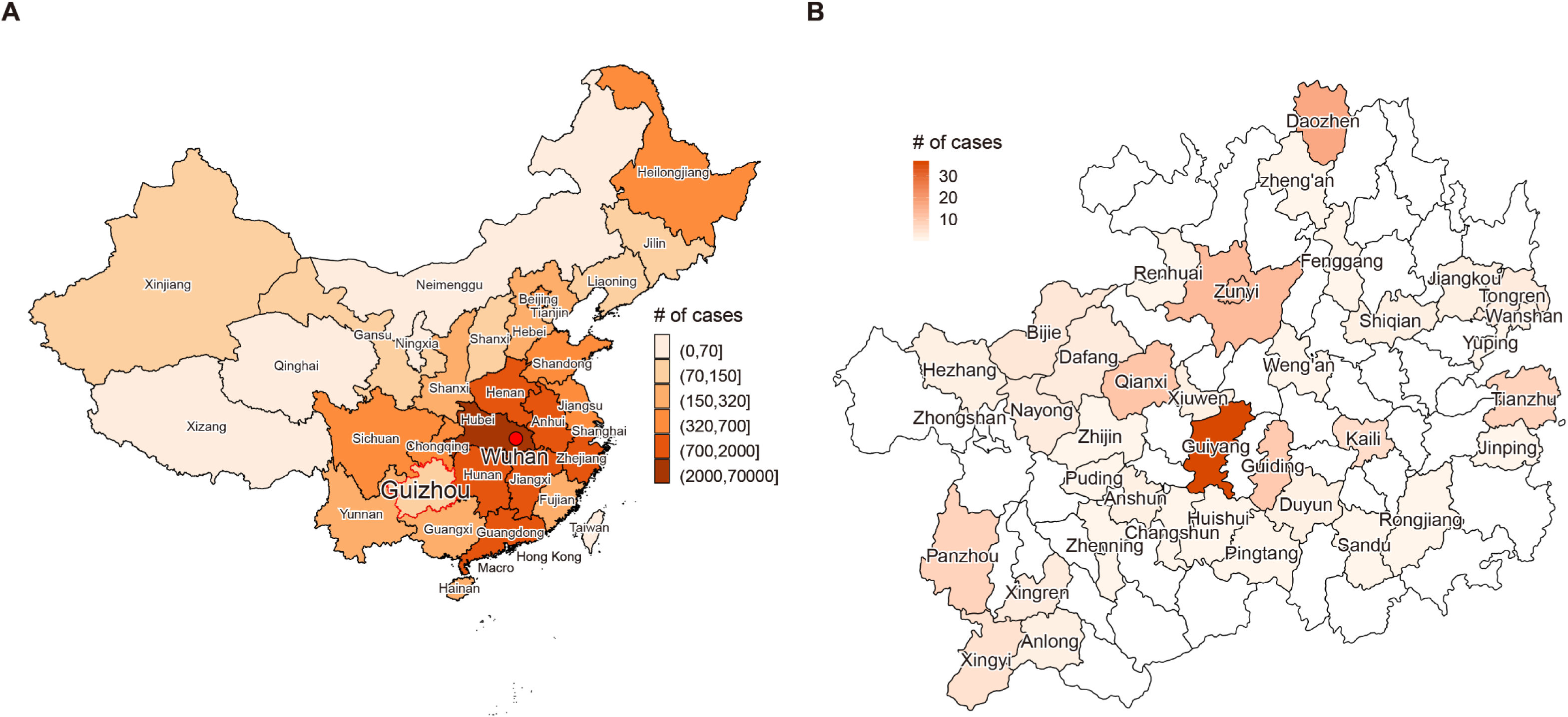
The geographic distribution of confirmed cases in China and Guizhou. **(A) Geographic distribution and** statistics of all confirmed cases in China as of February 16. Data was collected from National Health Committee^14^. In total 57,416 cases were confirmed throughout China, and 144 cases were in Guizhou. **(B) Geographic distribution and** statistics in Guizhou province as of February 16. In addition to confirmed cases with symptom, 18 cases without symptom were also included, leading to 162 cases shown in the figure. Guiyang, locating in the center of Guizhou, is the capital and largest city of the province, where 38 cases were reported. Throughout Guizhou province, 38 out of 88 administrative districts have reported confirm cases.

The first case was detected on January 21 and was officially confirmed by China CDC on January 23. It was reported by Guiyang (capital city of Guizhou) CDC. This case was a 51-year-old male, infected in Wuhan where he was there to do some business affairs. His onset date was on January 9 when he was still in Wuhan. After returning to Guizhou on January 14, he visited a local hospital for several times until January 16 when he was isolated by the Division of Infection. **Figure 2** shows the distribution of the onset date of all cases related to 2019-nCoV over time. It is notable that there was a decline of cases after February 7th, which is most likely attributed to the delay of reporting by the local hospital and laboratory confirmation.

**Figure 2.**
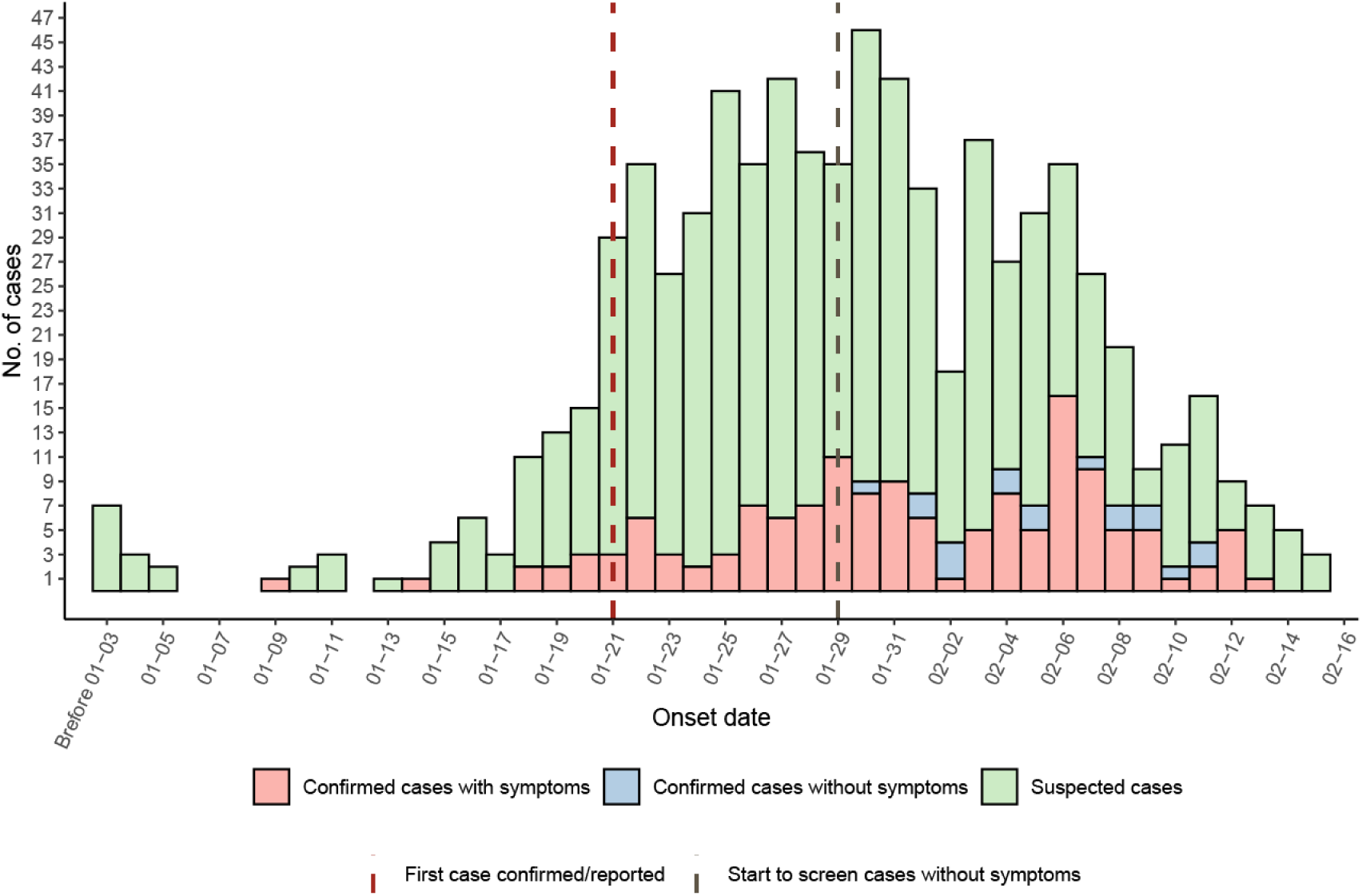
Distribution of the onset date of confirmed and suspicious cases of COVID-19 in Guizhou, China. The horizontal axis represents the onset date of confirmed cases or suspected cases. The onset data was defined as the self-reported first date to have symptoms related to 2019-nCoV and as the confirmation date for confirmed cases without symptoms. On January 21, the first case was officially confirmed and reported by Guiyang (capital city of Guizhou) CDC, whose onset date was on January 9th when the case was still in Wuhan. On January 29th, we started to identify and screen close contacts of confirmed cases and those had a positive result by real-time reverse-transcription– polymerase-chain-reaction (RT-PCR) yet without symptoms were labeled as “Confirmed cases without symptoms” in the figure. It is notable that there shows a decline of cases after February 12, which is most likely attributed to the delay of reporting by the local hospital and laboratory confirmation.

We included 93 confirmed cases with detailed exposure window and onset date information to estimate the incubation period (Fig. 3 A-B). The median incubation period was 8.06 days (95% confidence interval [CI]: 6.89 – 9.36). The 95% percentile was 21.90 days (95% CI: 18.21 – 25.45). Other percentiles are shown on **Table 2**. We obtained 57 observations from cluster cases and estimated the serial interval distribution (Fig. 3C) with the mean (±SD) of 6.37±4.15 days. The estimated basic reproduction number (R0) was 1.09 (95% CI: 0.94 – 1.26).

**Table 2.** Percentiles of incubation period

|  | Estimates | 95%CI Low | 95%CI High | Standard Error |
| --- | --- | --- | --- | --- |
| P2.5 | 2.455 | 1.716 | 3.565 | 0.485 |
| P5 | 2.972 | 2.15 | 4.162 | 0.523 |
| P25 | 5.358 | 4.302 | 6.625 | 0.611 |
| P50 | 8.069 | 6.897 | 9.362 | 0.636 |
| P75 | 12.152 | 10.64 | 13.514 | 0.733 |
| P95 | 21.904 | 18.219 | 25.452 | 1.833 |
| P97.5 | 26.523 | 21.356 | 31.775 | 2.635 |

-2\*Log Likelihood = 306.3

**Figure 3.**
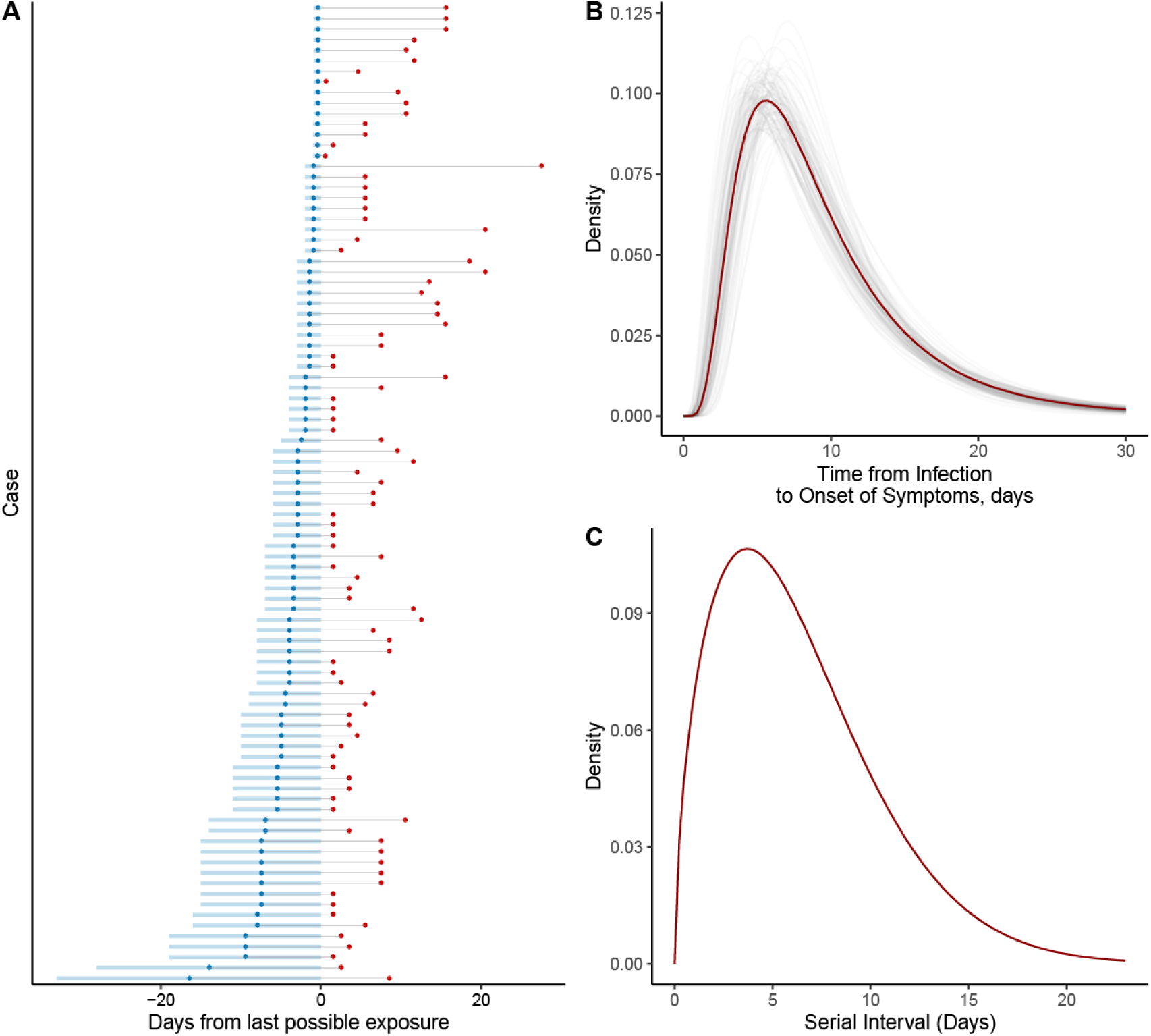
Key time-to-event distributions. **(A)** The time windows of exposure and onset points of symptoms for 93 cases (45 females, 48.38%; 48 males, 51.62%; median age 42, IQR: 27-55) that have detailed information of exposure and onset dates. Blue shaded regions indicate the exposure windows, and blue points represent the midpoint within the region. Exposures were defined as travel histories to Wuhan or close contact to other infectious individuals. **(B)** The estimated incubation period distribution using a log-normal model with 1,000 times bootstrap resamples. The estimated median incubation period in Guizhou is 8.06 days (95% CI: 6.89 – 9.36). It was estimated that approximately 2.5% of the individuals have symptoms in 2.45 days (95% CI: 1.71 – 3.56) after the infection, and 97.5% of that show symptoms within 26.52 days (95% CI: 21.35 – 31.77) after the infection. **(C)** The estimated serial interval distribution -- the duration between symptom onset of a primary case and symptom onset of its secondary cases -- fitted via the Weibull distribution, with a mean (±SD) of 6.37±4.15 days.

## Discussions

This study provides an initial analysis among epidemiologic characteristics and two typical transmission phenomena of COVID-19. Most index cases were exposed to Wuhan or confirmed cases related to Wuhan, but it is also necessary to prevent the community transmission of COVID-19. Measures such as community screening, isolation of close contacts, public transportation limitation, extended population of PCR testing, enhanced disinfection, and health education etc. could be taken. Our findings provide more epidemiologic data to this incident of COVID-19, which contribute to the further analysis and the control of this disease.

In addition to the evidence of the human-to-human transmission, it is likely that there exists infectiousness to some extend during the incubation period, and the presence of “super-spreaders”. The median incubation period was 8.2 days (95% CI: 7.9 – 9.5) in our study, which is longer than a recent report of 425 patients (8.2 days vs. 5.2 days), this may be a results of recall bias, during epidemiological investigation, we found that some cases actually have had shown mild symptoms days before the date they reported so as to their ignorance. We estimated the R0 as 1.09 (95% CI: 1.9 – 1.2), which means each patient can infect other 1.09 people on average. In commonly used infection models, when R0 > 1 the infection will be able to start spreading in a population, but not if R0 < 1.

However, it should be treated with caution that the basic reproduction number R0 was calculated under the assumptions that an infected individual but has no symptoms does not yet infect others, which assumes that all index cases should show symptoms before their secondary cases. However, as the investigation went on, Guizhou CDC found that there existed index cases who showed symptoms after that of their secondary cases. This may lead to that the estimated R0 was not robust as we do not hold enough evidence and information to the presence of asymptomatic carrier, those data were not included in the estimation model.

There are several limitations in our study. First, we could not collect all complete exposure and onset data from all confirmed cases, due to the recall bias and the lack of standardization on epidemiological investigation at the beginning of the incident. Second, some key data from asymptomatic carrier, especially the onset date or onset window, and left bound of exposure window from cases of Wuhan residents were unavailable, other statistical model may be used to fit such kind of censored data to estimate serial interval and R0 more accurately.

## Data Availability

The data that support the findings of this study are available from the corresponding author upon reasonable request.

## Author Bio

Kaike Ping acquires his Master degree of Biostatistics in Southern Medical University on 2017. Now he is a researcher at Center for Disease Control and Prevention of Guizhou, China, where he has been investigating respiratory diseases and their epidemiologic Characteristics. His research interests include disease distributions, time series analysis, survival analysis, and machine learning.

